# Operational but fragile: a mixed-methods assessment of outbreak Preparedness in Addis Ababa’s tertiary hospitals

**DOI:** 10.1101/2025.11.11.25340046

**Authors:** Mikiyas Alemu, Rebecca Shewaneka, Saron Ashenafi, Ashenafi Zelalem, Milkiyas Bedane, Ebenezer Fanta

## Abstract

**Background:** Emerging viral infections (EVIs) present a critical challenge to global health security, with tertiary hospitals serving as essential hubs for outbreak response. In low-resource settings like Ethiopia, these institutions face systemic vulnerabilities, yet a comprehensive assessment of their preparedness, encompassing both measurable capacities and underlying operational cultures, is lacking. This study aimed to evaluate the outbreak preparedness for EVIs for Addis Ababa’s tertiary hospitals.

**Methods:** A hospital-based, mixed-methods study was conducted between January and April 2025 public and private tertiary hospitals. Descriptive analysis was done for quantitative component gathered using structured observational checklist adapted from WHO and CDC tools, assessing ten core domains of preparedness, from coordination to logistics. Thematic analysis was performed on the qualitative data obtained through 21 in-depth interviews with purposively selected key informants, including hospital leadership, infection prevention personnel, and frontline clinicians.

**Results:** The median total preparedness score was 63.61/100, classified as ‘Operational Preparedness’. Laboratory capacity was the weakest domain (36.4/100), only 3.5% of staff were trained in outbreak response, and no hospital had negative-pressure isolation rooms. Only 30.4% of clinical staff had access to N95/N99 respirators. Qualitative interviews revealed that coordination was often improvised; as one emergency doctor stated, “Committees are formed immediately when such events occur, but there is no permanent structure.” Furthermore, the absence of proactive, disease-specific protocols led to reliance on generalized measures, delaying effective response.

**Conclusion/Implications:** While scoring at an operational level, the preparedness of these hospitals is fundamentally fragile and reactive. The infrastructural and training gaps are severely compounded by a non-proactive institutional culture and a lack of systematized protocols. Moving from reactivity to resilience requires substantial investment in laboratory infrastructure, permanent isolation facilities, and mandatory simulation training, coupled with institutionalizing leadership structures, developing disease-specific plans, and integrating psychosocial support for healthcare workers.

## Introduction

Emerging viral infections (EVIs) such as Ebola, Zika, and COVID-19 pose a growing threat to global health, particularly in low- and middle-income countries (LMICs) where healthcare systems are often under-resourced and unprepared (1, 2). Tertiary hospitals, which serve as critical hubs for specialized care and outbreak response, play a central role in emergency management, including patient isolation, coordination of care, and infection control. Their effectiveness, however, depends on factors such as infrastructure, trained personnel, and robust emergency management systems like Emergency Management Plans (EMPs), Emergency Operations Centers (EOCs), and Incident Management Systems (IMS) (3–5).

In Ethiopia, evidence indicates persistent challenges in hospital preparedness, including inadequate personal protective equipment (PPE), limited staff training, and weak coordination mechanisms (6). These issues are further exacerbated by systemic limitations such as fragmented health policies and insufficient funding (7). The COVID-19 pandemic has underscored the urgency of strengthening hospital preparedness, even in high-income settings, revealing global vulnerabilities in surge capacity, laboratory services, and infection control (8–10).

Despite their critical role, tertiary hospitals in LMICs like Ethiopia are often insufficiently studied in terms of outbreak preparedness. Existing research highlights common gaps in administrative readiness, emergency service organization, inter-hospital communication, and standardized documentation (11, 12). Hospitals also face ongoing logistical barriers, such as shortages in PPE and medical supplies, weak surveillance systems, and limited surge capacity (13, 14). This limits their ability to rapidly scale up care delivery and maintain essential services during crises.

This study focuses on tertiary hospitals in Addis Ababa and aims to assess their capacity to manage EVIs. It identifies gaps in preparedness across key domains such as infection prevention and control (IPC), staff readiness, logistics, and surveillance. In doing so, it contributes to global and local efforts to improve outbreak resilience, protect healthcare workers, and sustain critical health services during emergencies.

## Methods

### Study Design

A cross-sectional, mixed-methods study was conducted between January and April 2025. The quantitative component employed a descriptive cross-sectional design, while the qualitative component used in-depth interviews. This design allowed structured assessment of preparedness indicators alongside exploration of healthcare workers’ and administrators’ experiences with outbreak response. Integrating both methods facilitated triangulation of findings, enhancing validity, context, and interpretability.

### Study Setting and Population

Addis Ababa, the capital city of Ethiopia with a population exceeding four million, serves as the administrative and healthcare hub of the country. The Ethiopian health system is organized into a three-tier structure comprising Primary Health Care Units (PHCUs), general hospitals, and tertiary hospitals. Tertiary hospitals provide referral-level care and advanced emergency services and serve as focal points for national outbreak response.

Eight tertiary hospitals (seven public, one private) formed the sampling frame. Five hospitals participated in the study; two declined, and one psychiatric-focused hospital was excluded. The qualitative study population included healthcare workers and administrators directly or indirectly involved in outbreak preparedness. A total of 21 key informants were purposively selected across strata, including hospital leadership, infection prevention personnel, emergency coordinators, pharmacists, frontline clinical staff, and public health officers. Sampling continued until thematic saturation was reached.

### Survey Instruments

Quantitative data were collected using a structured observational checklist adapted from the WHO All-Hazards Hospital Emergency Response Checklist and the CDC Hospital Preparedness Self-Assessment Tool. The checklist was pilot-tested at a secondary hospital and refined prior to deployment. It covered ten core domains of hospital preparedness, including coordination, communication, resource management, infection prevention, and surveillance systems. Each item was scored as “Yes” or “No” and aligned with WHO capacity indicators.

Qualitative data were collected through semi-structured, in-depth interviews guided by a tool developed to explore participants’ experiences, perceptions, and challenges regarding outbreak preparedness. Interviews were conducted in confidential settings, audio-recorded with consent, and transcribed and translated into English. Relevant institutional documents, including standard operating procedures and emergency response protocols, were also reviewed to assess the presence, adequacy, and implementation of formal preparedness policies. Hospitals were anonymized and coded H1 through H5 to ensure confidentiality.

### Data Collection Procedures

The checklist and interview guides were administered onsite with the support of designated hospital coordinators. For qualitative data, interviews were conducted until no new themes emerged. All collected documents were reviewed systematically for completeness and relevance. Written Informed consent was obtained from all participants prior to interviews. Institutional permissions were also secured from each participating hospitals.

### Data Analysis

Quantitative data were entered into SPSS version 30.0, cleaned, and analyzed. Descriptive statistics, including frequencies, percentages, medians, and average deviations, were computed for hospital resources and preparedness indicators. Preparedness was scored on a six-point Likert scale from “No Preparedness” (1) to “Comprehensive Preparedness” (6). Domain-specific scores were normalized to a 0–100 percentage scale. Overall preparedness was calculated as the mean of median scores across the ten WHO domains. Hospitals were classified according to HEPSA (Hospital Emergency Preparedness Self-Assessment) levels based on total scores.

Qualitative data were analyzed thematically using OpenCode version 4.03. The process included open coding to label meaningful phrases, axial coding to group overlapping codes into categories, and selective coding to distill overarching themes. Themes were linked to study objectives and illustrated with representative quotations.

Integration of quantitative and qualitative data was performed through triangulation, allowing comparison of structured assessment results with interview insights to provide a comprehensive understanding of both measurable infrastructure and contextual factors affecting outbreak preparedness.

## Result

### Basic Characteristics and Preparedness Overview

Five tertiary hospitals, three federal, one state-run, and one private, were included in the study. A total of 21 key informants were purposively selected across professional groups, including administrators, IPC staff, emergency coordinators, clinicians, and public health officials, with a gender distribution of 11 females and 10 males.

While hospitals varied in size, the median staff count was 1,137, with 267 staffed ward beds. Negative-pressure isolation rooms were absent across all facilities. Surge capacity within 12 hours averaged 50 additional beds. (Table 1.)

**Table 1.** Hospital Bed Capacity and Surge Readiness Across Clinical Units for Tertiary Hospitals in Addis Ababa, 2025.

| Bed capacity | H1 | H2 | H3 | H4 | H5 | Total |
| --- | --- | --- | --- | --- | --- | --- |
|  | Median | Median | Median | Median | Median | Median |
| Staffed Beds: | 12 | 13 | 8 | 10 | 12 | 10 |
| ICU Medical |  |  |  |  |  |  |
| Staffed Beds: | 0 | 10 | 0 | 0 | 4 | 0 |
| ICU Pediatric |  |  |  |  |  |  |
| Staffed Beds: | 14 | 7 | 21 | 7 | 34 | 14 |
| NICU |  |  |  |  |  |  |
| Staffed Beds: | 160 | 200 | 267 | 714 | 410 | 267 |
| Wards |  |  |  |  |  |  |
| Staffed Beds: | 14 | 32 | 24 | 14 | 26 | 24 |
| ED |  |  |  |  |  |  |
| Beds with | 0 | 0 | 0 | 0 | 0 | 0 |
| Negative Air |  |  |  |  |  |  |
| Flow |  |  |  |  |  |  |
| Maximal case | 210 | 250 | 320 | 745 | 513 | 320 |
| admission |  |  |  |  |  |  |
| capacity (beds) |  |  |  |  |  |  |
| Additional beds | 20 | 70 | 50 | 80 | 45 | 50 |
| staffed/equipped |  |  |  |  |  |  |
| within 12 hours |  |  |  |  |  |  |

Based on the HEPSA scoring system, three hospitals were classified at Level 4 (Operational Preparedness), while the remaining two fell under Level 3 (Developing Preparedness). Overall institutional outbreak readiness scores ranged from 48.85 to 70.82, with a median total preparedness score across all five tertiary hospitals of 63.61, reflecting considerable variability in emergency response capacities (Table 2).

**Table 2.** Summary of Hospital Scores Across WHO Emergency Response Capacities for Tertiary Hospitals in Addis Ababa, 2025.

| WHO Capacity Area | H1 | H2 | H3 | H4 | H5 |  |
| --- | --- | --- | --- | --- | --- | --- |
|  | Median | Median | Median | Median | Median | Median |
| Coordination & Incident Management | 5.26 | 78.95 | 44.74 | 52.63 | 28.95 | 44.74 |
| Emergency Teams & Surge Staffing | 23.08 | 76.92 | 76.92 | 92.31 | 61.54 | 76.92 |
| Case Management & Clinical Operations | 70.73 | 73.17 | 68.29 | 73.17 | 75.61 | 73.17 |
| Surveillance & Early Warning Systems | 55.56 | 50.00 | 66.67 | 88.89 | 72.22 | 66.67 |
| Community Engagement & Risk Communication | 47.37 | 73.68 | 63.16 | 57.89 | 68.42 | 63.16 |
| Laboratory & Diagnostic Capacity | 72.73 | 27.27 | 54.55 | 36.36 | 36.36 | 36.36 |
| Operational Support & Logistics | 36.36 | 45.45 | 63.64 | 72.73 | 45.45 | 45.45 |
| Safety & Security | 39.13 | 65.22 | 60.87 | 73.91 | 30.43 | 60.87 |
| Coordination & Incident Management | 89.47 | 89.47 | 73.68 | 89.47 | 84.21 | 89.47 |
| Total Hospitals outbreak preparedness Score | 48.85 | 64.46 | 63.61 | 70.82 | 55.91 | 63.61 |

### Coordination and Incident Management

The coordination and incident management capacity showed marked variation across the five hospitals, reflected in a median preparedness score of 44.74. Only two facilities, one federal and one state-run, had documented Emergency Management Plans (EMPs). These EMPs included provisions for risk assessment, designated roles, and communication protocols, but coverage varied across domains (Figure 1). Three hospitals had established Emergency Operations Centers (EOCs), but only two were equipped with basic telephones, and just one had satellite phones or two-way radios. A fully functional backup communication system was reported by only one hospital, emphasizing gaps in the crisis communication infrastructure.

**Figure 1.**
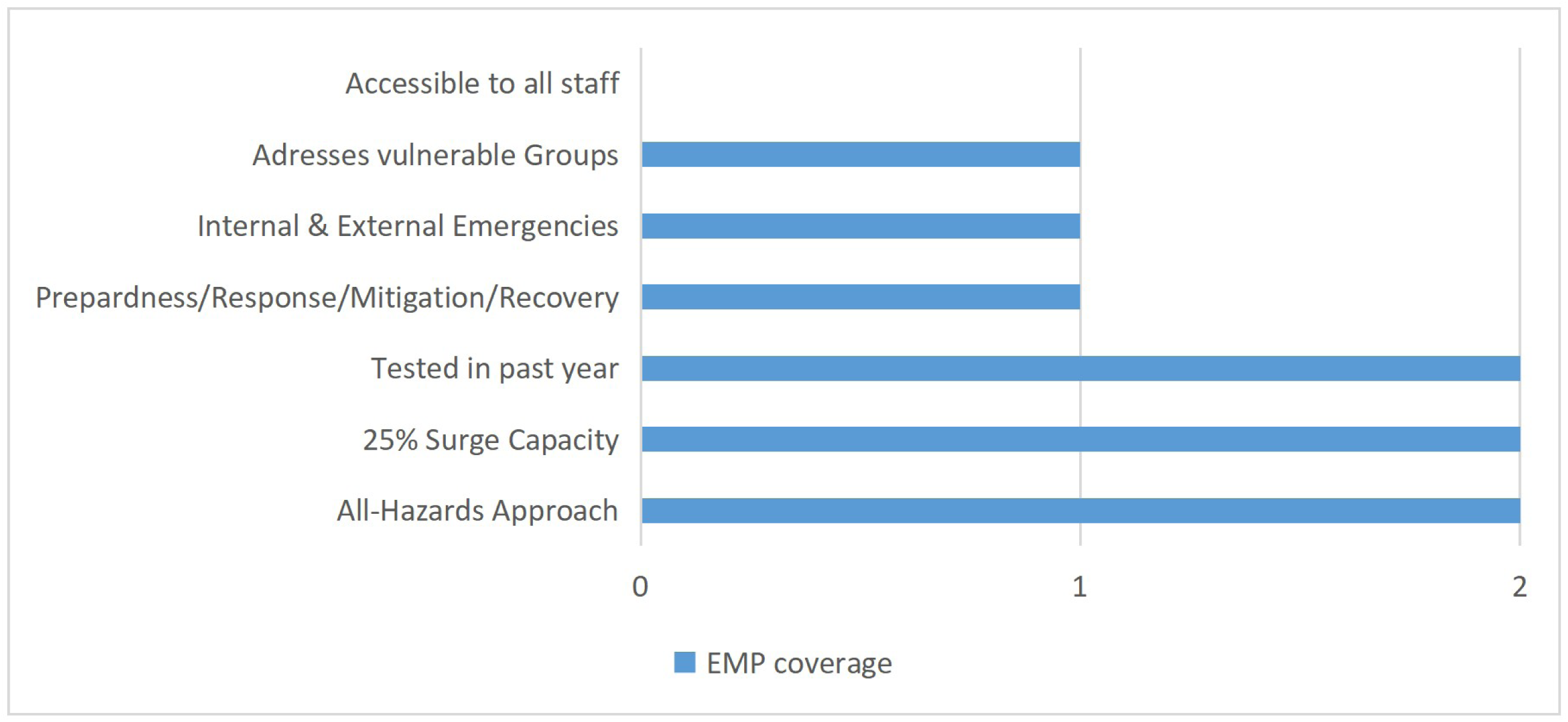
Coverage of key features in the Emergency Management Plans (EMPs) for tertiary hospitals in Addis Ababa, 2025.

Implementation of the Incident Command System (ICS) varied among hospitals. Only one facility reported conducting ICS exercises twice annually, while two others had conducted such exercises within the past six months. Four hospitals reported the presence of a multidisciplinary emergency response team, with clearly assigned roles and a designated Incident Commander. In these hospitals, all staff were aware of their roles and reporting locations upon ICS activation. The same four hospitals also reported functional communication infrastructure for the ICS command staff (Table 3).

**Table 3.**
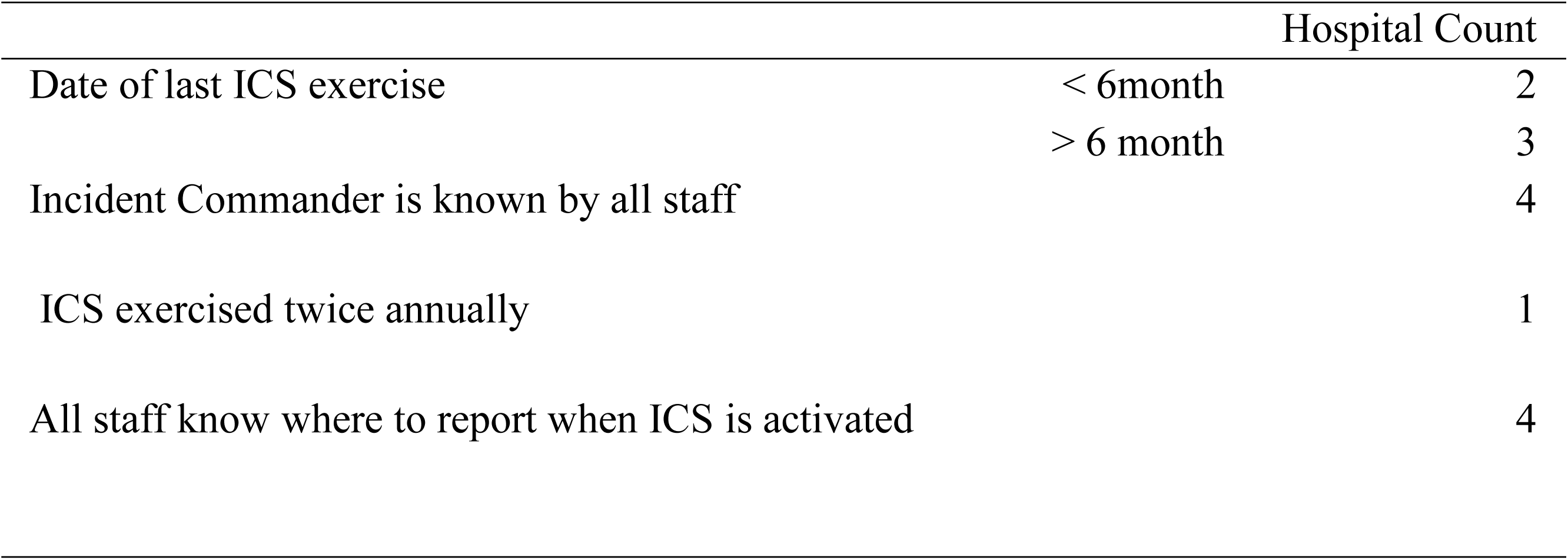
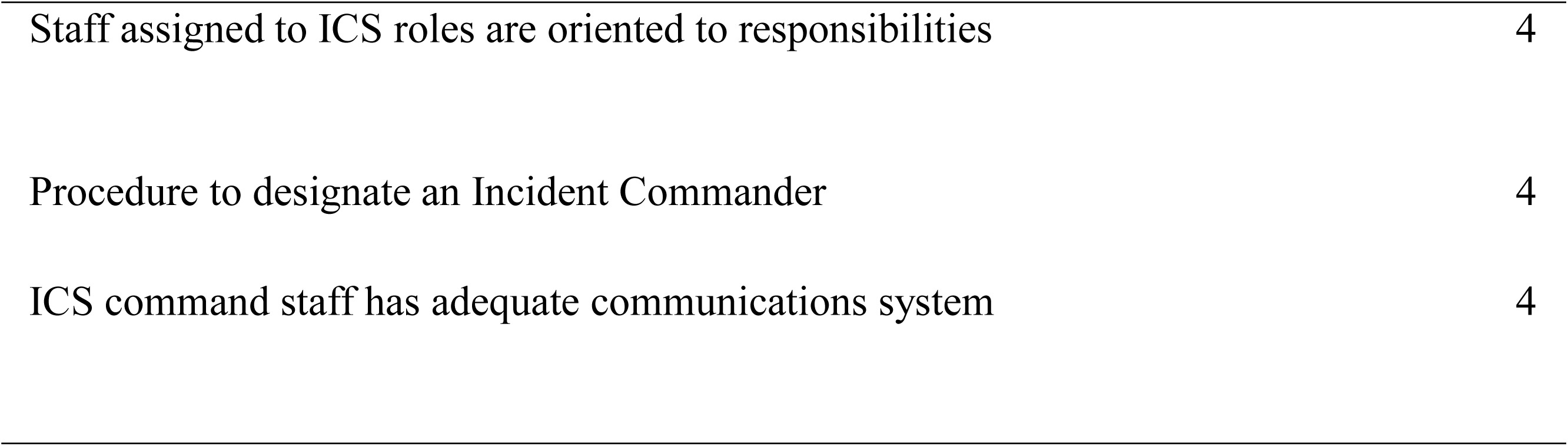
Implementation of ICS Components in Five Tertiary Hospitals in Addis Ababa for teritiary hospitals in Addis Ababa, 2025.

|  |  | Hospital Count |
| --- | --- | --- |
| Date of last ICS exercise | < 6month | 2 |
|  | > 6 month | 3 |
| Incident Commander is known by all staff |  | 4 |
| ICS exercised twice annually |  | 1 |
| All staff know where to report when ICS is activated |  | 4 |

|  |  |
| --- | --- |
| Staff assigned to ICS roles are oriented to responsibilities | 4 |
| Procedure to designate an Incident Commander | 4 |
| ICS command staff has adequate communications system | 4 |

Qualitative interviews reinforced these quantitative findings. Several respondents highlighted the reactive nature of emergency coordination in their facilities. An Emergency Doctor noted, *“Committees are formed immediately when such events occur, but there is no permanent structure,”* indicating that preparedness is often driven by crisis rather than planning. A Quality Officer added that, *“Coordination during COVID-19 was minimal due to the absence of a dedicated medical director,”* emphasizing leadership-related gaps. Another Hospital Coordinator remarked;

> “Well, our emergency response teams exist only on paper and are not functional in practice. Usually, preparedness efforts start only after cases are confirmed, so it ends up being reactive.”

### Emergency Medical Teams and Surge Staffing

Preparedness in this domain scored a median of 45.45, indicating significant limitations in staffing, training, and psychosocial preparedness. Only three hospitals had emergency medical teams capable of being activated during mass casualty events. The average percentage of staff trained in outbreak response protocols was low just 3.52% of total staff (range: 0% to 7.7%). Among clinical staff, the average training rate was slightly higher at 4.92% (range: 0% to 10.7%). Despite their critical frontline role, many emergency department personnel lacked consistent and comprehensive training.

Three hospitals conducted emergency drills across all shifts and departments, while the remaining two did so only occasionally. Four hospitals reported that their emergency departments participated in mass casualty simulations at least twice a year. However, drills were often limited to tabletop exercises rather than full-scale simulations.

Qualitative insights provided context for these numbers. An Infection Prevention Officer stated,

> “Training happens almost only during outbreaks, rather than being provided in a regular and structured manner. Although new staff are given brief orientations when they join, there is little confidence in their actual skills or preparedness for outbreak response.”

*Pointing* to a pattern of reactive training. Another Emergency Doctor highlighted the lack of realism in preparedness activities, saying, *“Simulation drills are rare and not taken seriously by most staff.”*

Human resource challenges were further emphasized in interviews. An Emergency and Critical Care Specialist remarked, *“Few staff are trained in advanced infection control,”* revealing insufficient depth in infection response capacity. Staff shortages and a lack of ongoing education were recurring themes.

Mental health and psychosocial support systems were weak across all facilities. Although all hospitals reported the availability of mental health services during and after outbreaks, only two had Critical Incident Stress Management (CISM) teams. While mental health professionals were included in Emergency Management Committees, only one hospital had staff members trained in crisis care and a system to assess the physical and psychological well-being of response teams.

These gaps were reflected in staff experiences during COVID-19. A frontline worker reported significant psychological strain: *“Healthcare workers faced burnout and exhaustion during COVID-19, impacting service delivery.”* No facility provided mandatory rest periods, nor was access to supportive conversations guaranteed. Only two hospitals offered reasonable duty hours, exposing most staff to a high risk of stress-related health issues.

### Case Management and Clinical Operations

This domain had a median preparedness score of 63.16, reflecting a moderate level of readiness for managing patients during outbreaks. All five hospitals reported the availability of essential emergency drug caches and used standardized triage protocols consistent with national EMS guidelines. However, triage systems were inconsistently applied: only three hospitals had clearly defined thresholds for triage activation, and significant limitations were observed in the physical infrastructure needed for surge triage and treatment (Figure 2).

**Figure 2.**
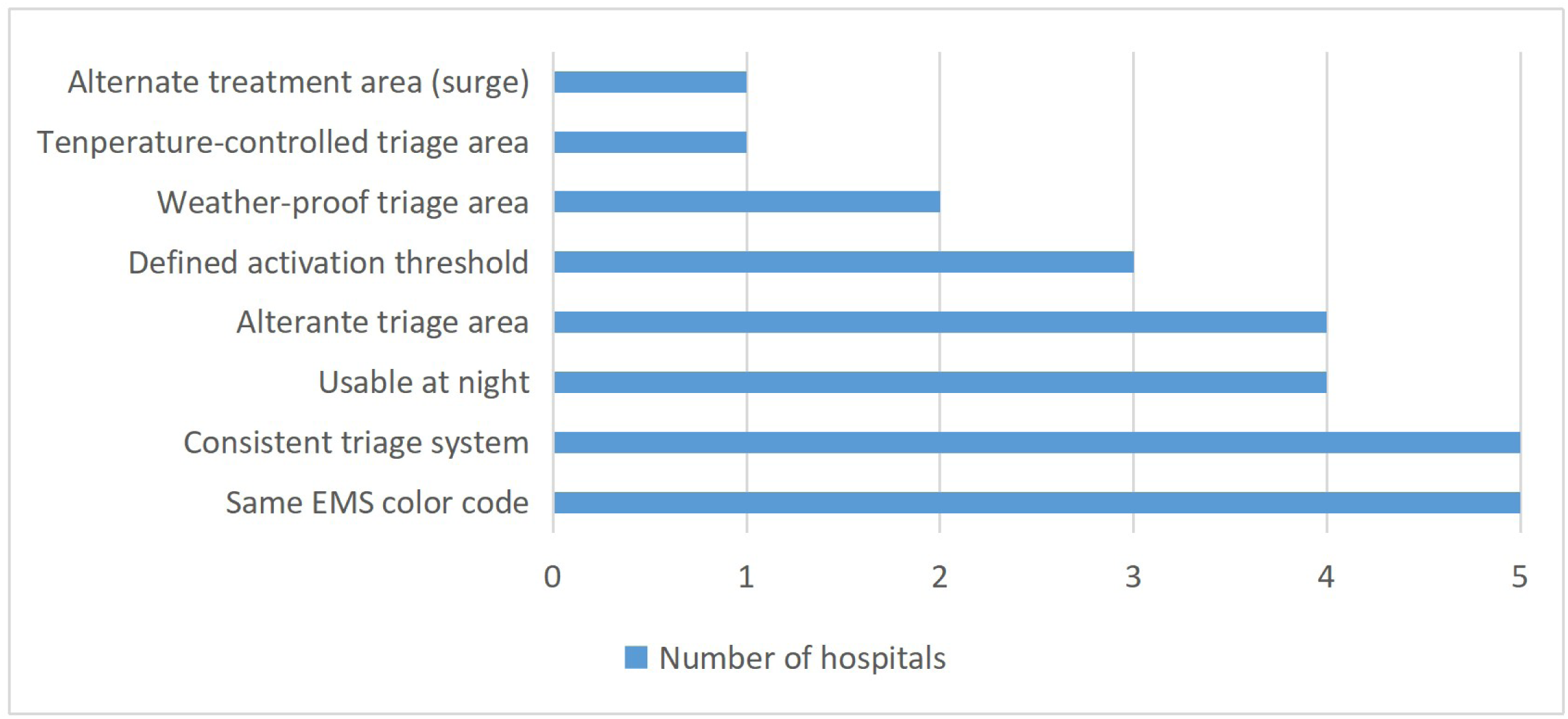
Coverage of Triage and Surge Treatment Features Across Hospitals for Tertiary Hospitals in Addis Ababa, 2025.

None of the hospitals had permanent alternate triage or isolation areas. During the COVID-19 pandemic, temporary solutions such as tents erected in hospital grounds were used to manage patient surges. One Emergency Doctor stated, *“During COVID, tents were set up in the hospital compound, but it’s not a permanent solution,”* highlighting the lack of durable infrastructure. Another emergency worker added, *“If an outbreak happens today, we would have to borrow space from other departments; there are no permanent isolation wings,”* underlining the risk posed by limited physical space for infectious case separation.

### Disease Identification and Surveillance

Hospitals demonstrated moderate capacity in disease detection and early warning, with a normalized preparedness score of 66.67. All facilities had structured surveillance systems, routinely reviewed acute febrile illness (AFI) and pulmonary diagnoses daily, and maintained timely coordination with public health authorities. Despite the strength of these surveillance frameworks, significant knowledge gaps among staff were reported regarding emerging and re-emerging viral infections.

While systems for daily case review and reporting existed, familiarity with specific high-consequence pathogens varied widely. Only one hospital provided staff with access to regularly updated case definitions or platforms containing disease-specific guidelines. Figure 3 provides a visual comparison of institutional familiarity with various emerging viral pathogens.

**Figure 3.**
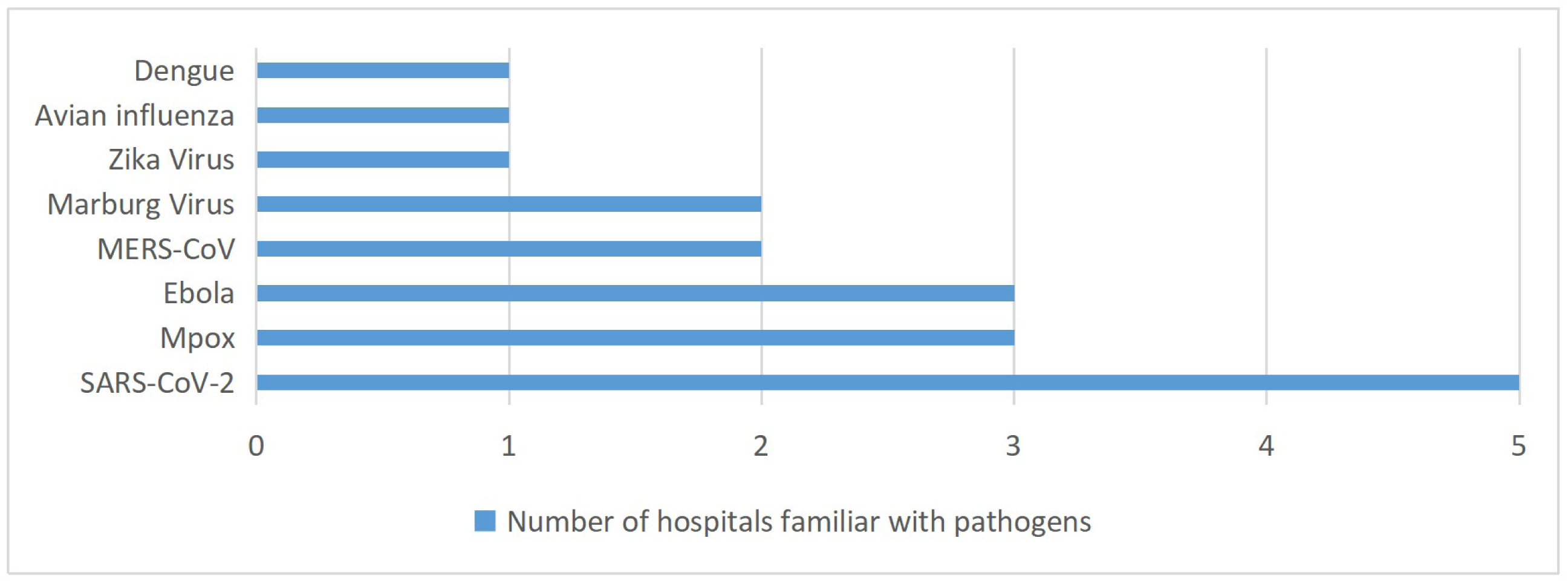
Hospital Familiarity with Emerging and Re-emerging Viral Pathogens for Tertiary Hospitals in Addis Ababa, 2025.

Qualitative responses echoed the quantitative gaps. One General Practitioner observed, *“Protocols exist, but when a new disease emerges like Mpox, we don’t have updated plans; we rely on general emergency measures.”* Another Public Health Emergency Officer emphasized the reactive nature of preparation efforts, stating, *“We only start preparations after we see confirmed cases.”* These insights highlight the systemic reliance on generalized emergency protocols, rather than proactive disease-specific planning.

### 6.7 Infection Prevention and Control (IPC)

Hospitals demonstrated variable readiness in IPC measures. While four of the five hospitals had personal protective equipment (PPE) available for clinical staff, only one facility had conducted proper fit testing for N95/N99 respirators. On average, 30.4% of clinical staff across the five hospitals were equipped with N95 or N99 respirators. This coverage ranged widely from 16% to 50%, indicating significant inequity in critical PPE distribution. In contrast, none of the non-clinical staff were reported to have any access to N95/N99 respirators. This highlights a potential vulnerability among ancillary hospital personnel during airborne infectious disease outbreaks. Clinical staff in all facilities had been trained in the use of PPE and immunization procedures. Four of the five hospitals had formal plans in place for antiviral procurement, and three had developed chemoprophylaxis protocols. Figure 4 summarizes the IPC and fatality management capacities across the facilities.

**Figure 4.**
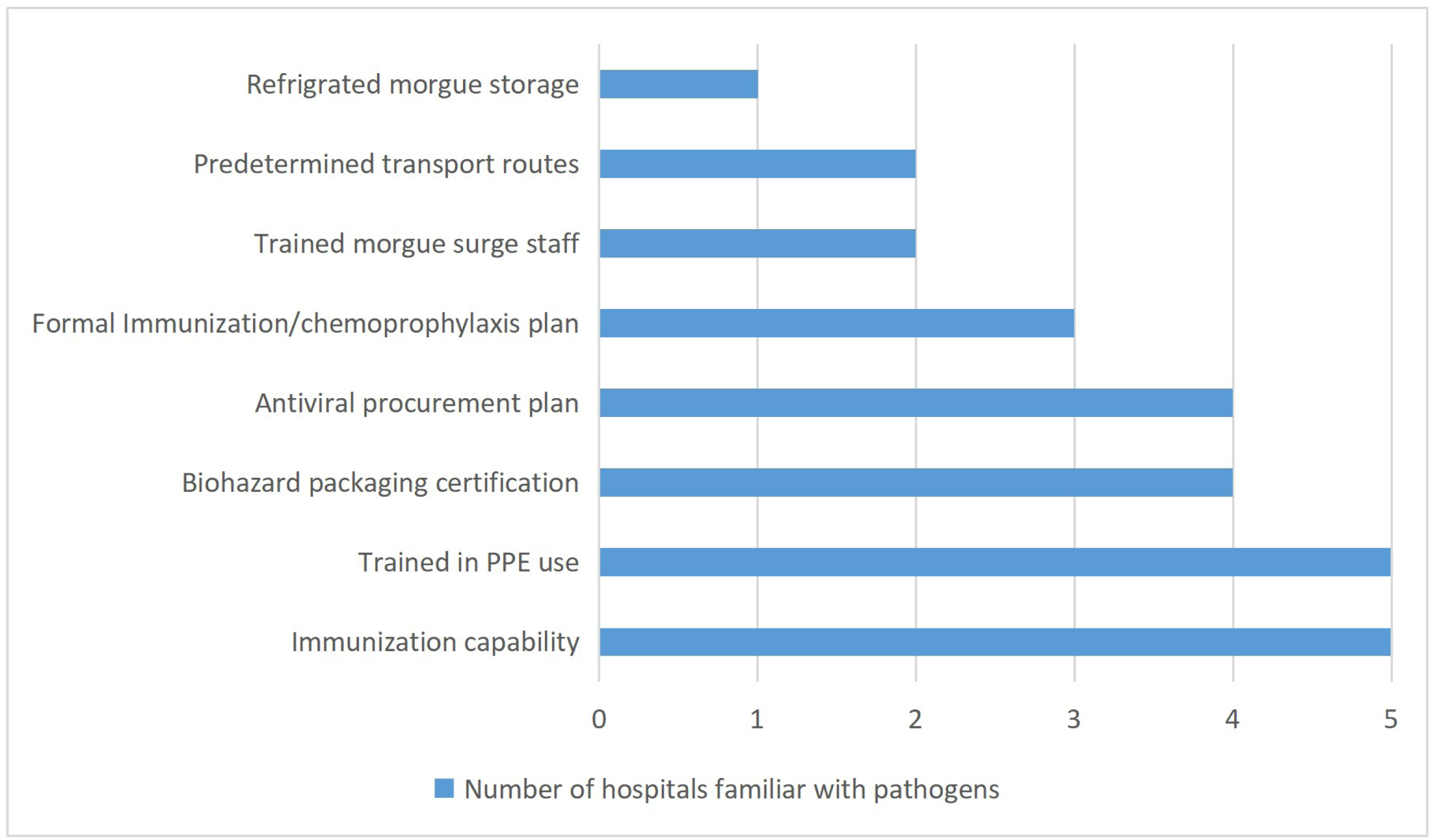
Infection Prevention, Fatality Management, and Biohazard Handling Capacities Across Hospitals for Tertiary Hospitals in Addis Ababa, 2025.

These findings were mirrored in qualitative interviews. Where a practice is driven more by supply limitations than by uniform policy or IPC protocol adherence. A General Practitioner explained,

> “We have an adequate supply of regular masks for routine use, but N95 masks are limited in availability. During periods of increased cases or surges, we prioritize these N95 masks specifically for staff working in high-risk areas to ensure their protection.”

### Risk Communication

Preparedness scores in this area were 60.87 for internal and external communication systems. All five hospitals demonstrated capacity to issue and receive emergency alerts and had mechanisms for notifying on- and off-duty staff. Contact databases were routinely updated and maintained for all clinical and non-clinical staff. Four hospitals had appointed Public Information Officers (PIOs), most of whom had pre-established relationships with media personnel.

Despite this formal structure, qualitative data suggested operational weaknesses in internal coordination. One Infection Prevention Officer noted, *“There is minimal coordination between administration and frontline health workers; only project-based communications occur.”*

Externally, hospitals lacked structured systems for engaging with the public during emergencies. Only a few facilities reported having designated information areas or crowd control plans. As a result, communication during emergencies was often uncoordinated and reactive. A nurse emphasized the implications of weak community engagement by stating,

> “Well, even if a hospital has all the plans and resources ready, it really doesn’t work without the community’s cooperation. People need to be involved and responsive; otherwise, the preparedness efforts just can’t function as intended.”

### 6.9 Laboratory and Diagnostics

Laboratory and diagnostic capacity was identified as one of the most critically weak domains, with a median preparedness score of 36.36, the lowest among all evaluated areas. Only two hospitals had in-house RT-PCR diagnostic equipment and operated at Biosafety Level 3 (BSL-3). The remaining three hospitals were limited to BSL-1 or BSL-2 environments and relied entirely on external laboratories for confirmatory PCR testing.

Laboratory surge readiness was extremely limited. Only one facility had identified backup lab staff and conducted specific training to handle surge testing capacity. All hospitals had protocols for reporting suspicious isolates to health authorities, indicating some degree of procedural compliance. However, only two hospitals had MOUs for resupplying critical reagents or for transferring workloads to alternate laboratories in case of capacity overflow.

Nevertheless, qualitative insights reveal a troubling decline in readiness since the peak of the COVID-19 pandemic and the waning institutional momentum for diagnostic preparedness post-pandemic. A Quality and Lab Officer reflected,

> “Whenever outbreaks are announced, we see a sudden spike in testing demand. During COVID-19, supplies tended to arrive quickly to meet the need, but at present, the level of readiness is much lower and harder to manage effectively.”

### Operational Support and Logistics

This domain demonstrated relatively strong preparedness, with a median score of 73.17. All five hospitals consistently stocked essential emergency medications, including epinephrine, corticosteroids, antibiotics, oxygen, saline, and nebulizers. ICU ventilators were available in each facility (range: 7–13; median: 10), while ER ventilators were limited (range: 0–2; average: 1). None of the hospitals had ventilators in general wards.

A critical logistics gap was noted in ventilator distribution and supply chain resilience. As one incident commander stated, *“Mechanical ventilators are lacking; if needed, we would have to procure them urgently, but there are shortages now.”*

All hospitals maintained secure storage for medical gases, and four had reserves for at least 72 hours. However, none had MOUs with secondary vendors for replenishment. Pharmacy readiness varied. While daily usage tracking was routine, only one hospital monitored antiviral spikes, and none tracked steroid use.

### Safety and Security

This component achieved the highest normalized preparedness score at 89.47, reflecting the robust systems in place for ensuring safety and continuity of operations during emergencies. All hospitals could enforce lockdowns and had security personnel at key entry points. Four hospitals had contingency plans to expand security staffing. Backup generators with three-day fuel supplies were functional in all hospitals, and critical services like labs and blood banks were prioritized during outages. While four of the hospitals conducted annual maintenance on power systems, ensuring operational resilience.

## Discussion

The overall assessment of tertiary hospitals in Addis Ababa revealed limitations in surge capacity, staffing, and isolation infrastructure, compromising their ability to manage emerging viral infections. Although these hospitals function as key referral centers, their capacity for medical and pediatric intensive care remains below international benchmarks for outbreak readiness (17). None of the facilities possessed negative-pressure isolation rooms, a foundational requirement for safe management of airborne pathogens such as SARS-CoV-2 and tuberculosis (15). Similar practice were seen in other low-income settings like Nigeria and Sierra Leone, where spatial adjustments remain the norm (18, 19). In Addis Ababa, studies have repeatedly highlighted the lack of specialized isolation facilities as a persistent gap (1). The variance in hospital ownership (federal, state, private) may partly explain inconsistencies in bed capacity and preparedness classification (three at operational and two at developing preparedness).

Coordination and incident management systems were generally weak. Only two hospitals had EMPs, and just one of these covered all phases of emergency response. Although some hospitals had EOCs, these were not systematically integrated into protocols. These findings suggest a low to moderate level of preparedness in incident management when evaluated against the WHO Checklist, which emphasizes institutionalized EMPs, regular drills, and accessibility of plans to all personnel (20). In countries with well-established emergency management systems, such structures are foundational to outbreak resilience (9). In Nigeria, ad hoc emergency committees dominate the response landscape, and structured EMPs remain rare, mirroring the findings of this study (3).

The proportion of hospital staff trained in outbreak response was notably low, with a median of 3.52% across the facilities. Simulation exercises were infrequently conducted, and in several hospitals, outbreak response roles were assigned without prior training, undermining institutional readiness for surge events. This reflects a low level of preparedness, especially when compared with international standards advocating routine cross-training, full-scale exercises, and surge staffing strategies (4, 21, 22). In sub-Saharan Africa, delayed and inadequate training during both the Ebola and COVID-19 responses contributed to increased morbidity and reduced healthcare workforce capacity (23).

Basic operational components, including standardized triage protocols and emergency drug stockpiles, were present across all hospitals. However, only one facility maintained a permanent, temperature-controlled alternate triage area, indicating limited investment in infrastructure for surge response. While hospitals indicated plans to maintain continuity of essential services during outbreaks, these strategies primarily relied on suspending elective procedures and reallocating resources. Although such measures align with global practices during health emergencies, they require deliberate planning to avoid compromising long-term population health. Evidence from the COVID-19 pandemic highlights that disruptions to routine care, particularly for individuals with chronic conditions, contributed to increased morbidity and mortality (24).

Hospitals maintained syndromic surveillance and routine reporting; however, gaps in staff knowledge and reliance on outdated clinical protocols limited timely detection of emerging infections. Only one facility had updated case definitions for specific EVIs, while others operated using general emergency guidelines. This disconnect between surveillance infrastructure and clinical expertise poses risks for delayed outbreak recognition. Comparable deficits in disease-specific knowledge have been reported in Nigeria and the Democratic Republic of Congo (25, 26). In contrast, hospitals in high-income settings routinely integrate updated protocols into clinical workflows, enabling rapid recognition and standardized response to novel pathogens (8, 14).

All hospitals had IPC Programs, but implementation varied, with limited PPE access; only 30.4% of clinical staff had N95/N99 masks, and none for non-clinical staff, and most lacked routine fit testing, compromising safety against airborne infections. During the COVID-19 pandemic, similar lapses in fit testing were linked to increased rates of occupational infection among healthcare workers (27). WHO recommends annual fit testing and inclusive PPE distribution for both clinical and non-clinical staff (15). In Belgium, these standards have been fully adopted, reflecting stronger IPC enforcement (28). In contrast, studies from Uganda and Sierra Leone report inconsistent IPC application and frequent exclusion of non-clinical staff from training and PPE allocation (29, 30). The exclusion of non-clinical staff from PPE allocation reflects systemic prioritization gaps and institutional oversights that diminish comprehensive outbreak preparedness.

Hospitals lacked structured public messaging and exhibited weak internal coordination, particularly during the COVID-19 response, despite having basic communication systems in place. Public health preparedness frameworks emphasize proactive communication, community risk education, and misinformation management as essential components (16). Literature from Nigeria and the Democratic Republic of Congo emphasizes the pivotal role of institutional trust and community engagement in effective disease control (9, 31). The absence of formal communication protocols and reliance on informal channels led to operational inefficiencies and undermined public trust, ultimately weakening coordinated emergency response efforts.

Laboratory and diagnostic capacity was the weakest domain across hospitals. Only two facilities maintained in-house RT-PCR testing with BSL-3 laboratories, while the rest relied on external services, resulting in delayed case confirmation. Similar constraints were reported in West African referral hospitals during the Ebola outbreak, where limited molecular diagnostics led to prolonged turnaround times (23). In contrast, tertiary hospitals in China operated on-site RT-PCR platforms with surge teams and reagent stockpiles, enabling timely diagnosis and improved outbreak control (11).

Operational support and logistics were generally adequate in supplying core resources such as emergency medications and oxygen. However, key gaps persisted, including limited ICU ventilator availability and the absence of tested backup vendor agreements both critical for sustaining responses during prolonged emergencies. WHO recommends redundancy in supply chains and a minimum 72-hour autonomous operating capacity to mitigate such risks (20, 21). Similar challenges have been documented globally in resource-limited settings, where advanced respiratory care infrastructure remains insufficient (17). These disparities are attributed to reliance on just-in-time procurement models and limited supplier diversification.

Safety and security were the strongest preparedness domain, with all hospitals demonstrating lockdown capacity, backup power systems, secure fuel reserves, and designated power continuity plans. These features align with WHO-recommended best practices, which emphasize lockdown capability, physical security, and uninterrupted power supply for essential services during emergencies (20, 32).

## Data Availability

All relevant data supporting the findings of this study are publicly available in the Mendeley Data repository at the following link: https://data.mendeley.com/datasets/d4ws6v3t58/1

## Notes

### Competing Interest Statement

The authors have declared no competing interest.

### Clinical Protocols

https://www.who.int/docs/default-source/documents/publications/hospital-emergency-response-checklist.pdf

https://calhospital.org/hospital-disaster-preparedness-self-assessment-tool/

### Funding Statement

None to Disclose.

### Author Declarations

The study was approved by the Institutional Review Board (IRB) of Myungsung Medical College (protocol PRO-310125, January 10, 2025) and the Addis Ababa Health Bureau IRB. Institutional permissions were obtained from all participating hospitals prior to data collection. two institutions declined participation and were therefore excluded. All data were anonymized before analysis using unique identifiers (AMR_ID) to ensure participant confidentiality.

